# Oxytocin enhances the recovery of eye-contact induced autonomic arousal: A treatment mechanism study with placebo-controlled design

**DOI:** 10.1101/2020.02.06.20020875

**Authors:** Nicky Daniels, Jellina Prinsen, Javier R. Soriano, Kaat Alaerts

## Abstract

The neuropeptide oxytocin (OT) is suggested to exert a pivotal role in a variety of complex human behaviors, including trust, attachment, social perception and fear-regulation. Previous studies have demonstrated that intranasal administration of OT reduces subjective and neuroendocrine stress responses and dampens amygdala reactivity. Moreover, OT has been proposed to modulate activity of the autonomic nervous system.

Here, we conducted a double-blind, placebo-controlled study with 56 men, to investigate whether a single-dose of OT (24 IU) modulates sympathetic autonomic arousal upon live dyadic gaze interactions. To do so, electro-dermal recordings of skin conductance were performed during the engagement of eye contact with a live model in a naturalistic two-person social context.

In accordance to prior research, direct eye gaze elicited a significant enhancement in skin conductance responses, but OT did not specifically enhance or dampen the overall magnitude (amplitude) of the autonomic arousal response. Administration of OT did facilitate the recovery of skin conductance responses back to baseline (increased steepness of recovery slope), indicating a role of OT in restoring homeostatic balance. Notably, the treatment-effect on autonomic recovery was most prominent in participants with low self-reported social responsiveness and high attachment avoidance, indicating that person-dependent factors play a pivotal role in determining OT treatment-responses. Behaviorally, OT significantly reduced self-reported feelings of tension and (at trend-level) worrying about how one presents oneself.

Together, these observations add further evidence to a role of OT in reducing subjective and autonomic stress responses, primarily by facilitating restoration of homeostatic balance after (social) stress-induced perturbation.

## Introduction

The neuropeptide oxytocin (OT) has been implicated to exert a pivotal role in a variety of complex human behaviors, including trust, attachment, social perception and fear regulation. The neuropeptide is synthesized in the magnocellular neurons of the supraoptic and paraventricular nuclei of the hypothalamus and through projections to the posterior pituitary gland, it is released to the bloodstream to act as a hormone and impact bodily functions. At the level of the brain, OT acts as an important neurotransmitter/ neuromodulator of a wide-range of complex social behaviors by impacting distinct subcortical and cortical brain structures through direct axonal projections (fast, focal) or continuous diffusion (slow, global) (Stoop, 2012).

Mechanistically, OT’s impact on human brain function has been proposed to involve a bottom-up anxiolytic effect to facilitate social approach behavior; and a top-down social salience effect to facilitate attention to, and perception of social signals (Ma, Shamay-Tsoory, Han, & Zink, 2016; Shamay-Tsoory & Abu-Akel, 2016). Several intranasal OT (IN-OT) administration studies have consistently shown that OT facilitates the recognition of emotional expressions from faces (Shahrestani, Kemp, & Guastella, 2013; van IJzendoorn & Bakermans-Kranenburg, 2012). Several studies also showed that OT increases the gaze time towards the eye region of faces both from static pictures (Guastella, Mitchell, & Dadds, 2008) (Eckstein et al., 2019) and during real-time interactions (Auyeung et al., 2015; Hall, Lightbody, McCarthy, Parker, & Reiss, 2012) (although see (Hubble et al., 2017; Lischke et al., 2012; Prinsen, Brams, & Alaerts, 2018) for contradictory results). With respect to OT’s anxiolytic role, several studies have demonstrated decreases in subjective reports of anxiety and reductions in neuroendocrine stress responses (cortisol) after IN-OT (Heinrichs, Baumgartner, Kirschbaum, & Ehlert, 2003; Meinlschmidt & Heim, 2007; Riem, Kunst, Bekker, Fallon, & Kupper, 2019). The diverse neuromodulatory effects of OT have been attributed to be mediated at least partly by OT’s influence on amygdala activation, a key neural structure of both fear-processing and (social) salience networks. For example, the seminal study by Kirsch et al. (2005) demonstrated that IN-OT induced and attenuation in amygdala activity during the processing of fearful or threatening visual images and a reduction in the coupling between amygdala and brainstem regions involved in autonomic functions (Kirsch et al., 2005).

Also, initial studies examining the autonomic effects of OT demonstrated an attenuating effect of OT on electrodermal or skin conductance (SC), a measure of physiological arousal driven solely by the sympathetic branch of the autonomic nervous system. For example, de Oliveira et al. (2012) demonstrated that IN-OT decreased subjective anticipatory anxiety as well as tonic autonomic arousal (skin conductance levels (SCL)) before and during a simulated Public Speaking Test (de Oliveira, Zuardi, Graeff, Queiroz, & Crippa, 2012). Likewise, Vietnam veterans with post-traumatic stress disorder showed lowest mean autonomic arousal (SCL) during personal combat imagery after receiving IN-OT (Pitman, Orr, & Lasko, 1993). Also associations between plasma OT levels and habituation in skin conductance responses (SCRs) during a trust game have been demonstrated (Keri & Kiss, 2011). In contrast to these studies, Gamer et al. (2012) investigated the effect of OT on sympathetic (SCRs) and parasympathetic (heart rate changes) function during a facial emotion classification task, but only identified an effect of IN-OT on heart rate changes, not on sympathetically driven autonomic arousal responses (Gamer & Buchel, 2012). Further, in a series of studies, Eckstein et al. (2015, 2016) demonstrated that while IN-OT enhanced the decline in SCRs during late phases of fear extinction (Eckstein et al., 2015), IN-OT also strengthened Pavlovian fear conditioning during the acquisition phase by enhancing SCRs to fear-associated stimuli (Eckstein et al., 2016). As noted by the authors, these findings speak against a strong account of OT having purely anxiolytic-like effects, but instead suggest that OT enables a rapid and flexible adaptation for the detection and processing of potential fear signals (Eckstein et al., 2016).

The notion of a complex dualistic (anxiolytic-anxiogenic) role of OT has been highlighted before and is generally corroborated by the identification of both down- and up-regulation of amygdala activity after IN-OT, as reviewed in recent meta-analytic reports of pharmaco-imaging studies (Grace, Rossell, Heinrichs, Kordsachia, & Labuschagne, 2018; Wang, Yan, Li, & Ma, 2017). As elegantly proposed by MacDonald et al. (2014), the seemingly dual action of OT on central and autonomic function during processing of social signals may stem from the ambiguous nature of social signals themselves, which in particular contexts, may signal reward, safety and pleasure (e.g., mother-infant bond), but in others, may signal competition, threat and stress (MacDonald & Feifel, 2014). Also evolutionary adaptations in perceiving and processing social cues, particularly eye-region related signals need to be considered to understand their ambiguous nature. For many animal species, a pair of watching eyes forms a highly salient ‘threat’ cue that reliably activates fear processing circuits and the sympathetic branch of the autonomic system, triggering defensive behavior or fighting responses (i.e. ‘flight or fight’), as they most likely signal the presence of a dominant conspecific or predator (Emery, 2000; Skuse, Morris, & Lawrence, 2003). In humans however, who have evolved to navigate complex social environments, the reflexive arousal responses evoked by direct gaze have been adapted in order to be used for other survival-related purposes, such as facilitating cooperative interaction and the formation of bonds (e.g. maternal-infant bonds), as well as signaling one’s own and inferring others’ emotional states and social intentions (Grossmann, 2017). While previous research consistently showed that direct eye contact forms a salient cue for inducing reliable autonomic arousal responses (i.e., higher SCRs upon direct, reciprocated gaze, compared to unreciprocated gaze (averted gaze or closed eyes)) (Helminen, Kaasinen, & Hietanen, 2011; Hietanen, Leppanen, Peltola, Linna-Aho, & Ruuhiala, 2008; Prinsen & Alaerts, 2019; Prinsen, Deschepper, Maes, & Alaerts, 2019), it remains currently unexplored whether and how IN-OT modulates autonomic arousal responses during the processing of direct eye gaze cues. As outlined by Skuse et al. (2003), enhanced autonomic arousal upon perceived eye gaze is driven predominantly by amygdala-centered subcortical circuits, but depending on the context of the social encounter (e.g. familiarity), amygdala-driven arousal upon eye contact can be modulated through neocortical top-down regulation (Skuse et al., 2003). Importantly, excessive arousal responses upon direct eye contact have been identified in individuals with social anxiety disorders (Myllyneva, Ranta, & Hietanen, 2015) and children with autism spectrum disorder (ASD; (Joseph, Ehrman, McNally, & Keehn, 2008; Kaartinen et al., 2012; Kylliainen & Hietanen, 2006), indicating that ineffective regulation of eye-contact induced autonomic responses may form an important hallmark of neuropsychiatric conditions in which social function is compromised.

The present study adopted a double-blind, randomized, placebo-controlled, between-subject design to examine the effect of IN-OT on autonomic arousal (SCRs) upon presentation of direct eye contact. In line with prior research, the eye contact stimuli were conveyed by a live model in a naturalistic two-person social context, which has been shown to elicit stronger arousal effects as compared to video recordings or pictures of faces (Hietanen et al., 2008; Ponkanen, Peltola, & Hietanen, 2011; Prinsen & Alaerts, 2019). Considering prior reports of attenuated autonomic arousal after IN-OT (e.g. reduced SCL during a public speaking task (de Oliveira et al., 2012)) and in support of the anxiolytic, stress-reducing role attributed to OT, we generally hypothesized OT to *reduce* autonomic arousal responses upon direct eye contact. However, considering that increased sympathetic drive (arousal) is linked to attention allocation and heightened perceptual processing, the possibility was not ruled out that IN-OT would induce an *increase* in SCRs upon direct eye contact, which would be indicative of a predominant effect of OT on top-down enhancement of social salience. Also subjective reports of experienced arousal, valence and self-consciousness (during the presentation of direct eye contact) were obtained to explore whether OT-induced changes in SC autonomic arousal are paralleled by changes in subjective experiences. Furthermore, considering recent notions that OT’s treatment responses are susceptibly modulated by person-dependent factors (Bartz et al., 2015; Bartz, Zaki, Bolger, & Ochsner, 2011), we explored whether inter-individual differences in self-reported social functioning (Social Responsiveness Scale) (Constantino et al., 2003) and attachment style (State Adult Attachment Measure, assessing dimensions of attachment security, avoidance and anxiety) (Gillath, Hart, Noftle, & Stockdale, 2009) potentially modulated identified treatment-responses.

## Experimental procedures

### Participants

A total of 56 healthy adults (all right-handed, Dutch speaking males) were recruited to participate in this double-blind, randomized, placebo-controlled study with parallel design. Due to technical difficulties, electro-dermal (skin conductance) recordings were not available for six participants (3 oxytocin; 3 placebo), such that analyses were performed on a total of 50 participants (25 oxytocin, mean age 22.0 ± 3.20 years; 25 placebo, 22.4 ± 3.42 years). Only male participants were included to avoid the potential confounding gender-dependent effects of OT (Zink & Meyer-Lindenberg, 2012). Participants abstained from alcohol and caffeine 24-hours before testing. Written informed consent was obtained from all participants prior to the study. Consent forms and study design were approved by the UZ / KU Leuven Ethics Committee for Biomedical Research in accordance to The Code of Ethics of the World Medical Association (Declaration of Helsinki). The study procedure was pre-registered at ClinicalTrials.gov (ClinicalTrials.gov Identifier: NCT03272321).

### Nasal spray administration

Participants were randomly assigned to receive a single dose of oxytocin (OT) or placebo (PL) based on a computer-generated randomized order. Except for the manager of randomization and masking of drug administration, all participants and research staff conducting the study were blind to treatment allocation. In correspondence with previous studies (see (Guastella & MacLeod, 2012) for a review), participants received 24 IU of OT (Syntocinon®, Sigma-tau) or placebo containing a saline natriumchloride solution. Participants received three puffs per nostril in an alternating fashion with each puff containing 4 IU. Participants were asked to first remove air present in the nasal spray by pumping the spray until a fine mist was observed. For inhalation of the spray, they were instructed to take a deep breath through the nose and to tilt their head slightly backwards during nasal administration in order to minimize gravitational loss of the spray. All participants were monitored onsite until approximately one hour after nasal spray administration and were then screened for potential side effects. Overall, only minimal, non-treatment specific side effects were reported, although note that a larger proportion of participants receiving the OT spray reported a (mild) headache (OT group: 6 out of 25; PL group: 1 out of 25) (see **Supplementary Table 1**). Finally, at the end of the experimental session, participants were asked if they thought they had received OT or PL or whether they were uncertain about the received spray. The majority of participants reported to be uncertain about the received spray (OT: 48%; PL: 48%) or thought they had received the PL nasal spray (OT: 32%; PL: 40%). The proportion of participants that believed they had received the OT treatment was not significantly larger in the actual OT group (20%), compared to the PL group (12%) (Pearson Chi-square 0.60; p=.44).

### Experimental procedure and stimuli

Prior to the electro-dermal recordings and nasal spray administrations, participants completed the Social Responsiveness Scale (SRS) (Constantino et al., 2003) and the State Adult Attachment Measure (SAAM) (Gillath et al., 2009).

The 64-item Dutch adult self-report version of the SRS comprises four subscales examining social communication, social awareness, social motivation and rigidity/repetitiveness, using a four-point Likert-scale.

The SAAM comprises three subscales examining attachment security (e.g., *“I feel like I have someone to rely on”*) (7 items); attachment anxiety (e.g., *“I feel a strong need to be unconditionally loved right now”*) (7 items); and attachment avoidance (e.g., “*If someone tried to get close to me, I would try to keep my distance”*) (7 items) using a seven-point Likert-scale.

Participants also completed the 32-item short version of the Profile of Mood States (POMS) questionnaire (McNair & Lorr, 1964) at the start (before nasal spray administration) and end of the experimental session (post nasal spray administration) as a measure of transient mood levels in five domains: tension (6 items), depression (8 items), anger (7 items), fatigue (6 items) and vigor (5 items). The POMS comprises emotional adjectives for which subjects need to indicate to what extent the word fits their current mood using a five-point Likert scale from “not at all” to “very well”. Note that reports of mood states were not recorded for three participants (all placebo), such that analyses were performed on a total of 28 OT and 25 PL participants.

After completion of the questionnaires, participants were seated in front of a 20 × 30 cm voltage-sensitive liquid crystal (LC) shutter screen (DreamGlass Group, Spain) attached to a black frame separating the participant from the live model (similar setup as used in (Prinsen et al., 2019)). Participants were instructed to observe and pay close attention to the stimuli presented through the screen. The presented stimuli comprised the face of a live female model sitting with eyes open and gazing towards the participant (i.e. showing direct gaze and engaging in mutual eye contact) or sitting with eyes closed. During stimulus presentation, the model bore a neutral expression and kept her face as motionless as possible and tried to avoid eye blinks. Two female experimenters, trained to act similarly towards the participants, served as models. The models were unknown to the participants and only had a brief standardized interaction with the participant before stimuli presentation. For each participant, the same model was presented pre- and post-nasal spray administration. The participant was seated 60 cm in front of the shutter at the same height as the model’s face with an overall distance between the participant and the model of 110 cm. Signal 4.11 software was used to trigger the LC window to shift from an opaque to a transparent state. Each stimulus (eyes open, eyes closed) was presented 10 times with duration of 5 sec in a semi-random sequence (no more than three consecutive trials of the same type). During all trials, electro-dermal recordings (skin conductance) were acquired. The inter-stimulus interval (ISI) varied with a minimum of 20 seconds to allow skin conductance responses to return to baseline.

### Subjective ratings of eye contact

After the presentation of the live gaze stimuli, participants completed three items of the Situational Self-Awareness Scale (SSAS) (Govern & Marsch, 2001) (Dutch translation) to assess public self-awareness in response to seeing a face with direct gaze *(“I was concerned about the way I present myself”, “I was self-conscious about the way I look”*, and *“I was concerned about what other people think of me”*). Responses were obtained on a visual analog scale from 1 = “completely agree” to 7 = “completely disagree”. The answers were reversed scored such that higher scores correspond with more self-awareness. Further, two items of the Self-Assessment Manikin (SAM) (Bradley & Lang, 1994) were adopted to assess the participants’ arousal and affective valence to seeing a face with direct gaze (9-point scale; 1 = calm/pleasant, 9 = arousing/unpleasant). Note that subjective ratings were not recorded for two participants (1 OT; 1 placebo), such that analyses were performed on a total of 27 OT and 27 PL participants.

### Electro-dermal recordings and data handling

The Nexus-32 multimodal acquisition system and BioTrace+ software (version 2015a, Mind Media, The Netherlands) were used to collect electro-dermal recordings (skin conductance). Other neurophysiological recordings (e.g. electroencephalography) were performed simultaneously, but these measures are not part of the current report. Two Ag/AgCl Velcro snap-on electrodes were attached to the palmar surface of the distal phalanxes of the index and middle fingers on the participant’s non-dominant hand to record stimuli-specific skin conductance responses (SCR) with a sampling rate of 128 Hz. Before electrode attachment, the skin was prepared with electrode paste (0.5% saline in a neutral base).

The SCR-amplitude was defined as the maximum change in amplitude relative to baseline during the five seconds after stimulus onset. The baseline was determined as the average of the two seconds right before stimulus onset. Maximum change from baseline amplitude scores below 0 µS were set to zero. SCR-amplitude data, were averaged separately for each condition and each participant to obtain an average score of the magnitude of the electro-dermal SCR (Myllyneva et al., 2015).

Stimuli-specific SCRs are typically characterized by a rise from initial level to a peak within 1-4 s after stimulus onset, followed by a recovery period of approximately 20 – 30 s in which the electro-dermal signal declines back to baseline (recovery point). In order to characterize the steepness of the recovery, SCR-slopes were computed as the absolute difference in SCR between the peak and recovery point, divided by the recovery time (i.e., Δ SCR_peak to recovery_ / Δ time_peak to recovery_); measured in µS/ sec. SCR recovery slopes were set to zero for trials in which no recovery was reached. Higher SCR slope values indicate steeper decline (faster recovery). Note that for two participants (both placebo), extreme outlier SCR-slope scores were identified (exceeding Q3 ± 5*(Q3-Q1), with Q1 and Q3 being the first and third quartile) and recoded to the mean (results are reported with and without recoding).

### Statistical Analysis

SCR-amplitudes and SCR-slopes data were subjected to a repeated-measures ANOVA analysis with the between-subject factor ‘treatment’ (OT, PL) and the within-subject factors ‘condition’ (eyes open, eyes closed) and ‘time’ (pre, post). Reports of mood states (five states) and subjective ratings of eye contact (self-awareness, pleasantness, arousal) were also subjected to repeated-measures ANOVA analyses with the between-subject factor ‘treatment’ (OT, PL) and the within-subject factor ‘time’ (pre, post). For all analyses, the significance level was set at p < .05. All statistics were executed with Statistica 13 (StatSoft. Inc. Tulsa, USA).

## Results

### Effect of IN-OT on skin conductance

#### Skin conductance response - amplitude

In accordance to prior literature (Helminen et al., 2011; Hietanen et al., 2008; Prinsen & Alaerts, 2019; Prinsen et al., 2019), ANOVA analysis of the SCR-amplitude data revealed a main effect of ‘condition’ (*F*(1, 48)= 12.48; *p*= .0009), indicating that across sessions and treatment groups, SCR-amplitudes were higher during the direct eye gaze condition, compared to the eyes closed condition (**Figure 1A**). Also a main effect of ‘time’ was revealed (*F*(1, 48)= 8.97; *p*= .004), indicating that in both treatment groups, SCR-amplitudes significantly declined from the pre- to the post session. Also a tentative ‘time x condition’ interaction effect was revealed (*F*(1, 48)= 3.51; *p*= .067), indicating that the condition effect (eyes open > eyes closed) was most pronounced at the pre-session (*t*(49)= 3.19; *p*= .0025), compared to the post-session (*t*(49)= 2.00; *p*= .052). The ‘treatment x time’ interaction was not significant (*F*(1, 48) < 0.01; *p*= .99) (nor any of the other interactions with the factor ‘treatment’: *p*> .70), indicating that pre-to-post changes in SCR-amplitudes were not significantly different between the OT and PL group. **Table 1** lists the pre-to-post change scores separately for each treatment group, as well as the treatment x time interaction effect.

**Figure 1.**
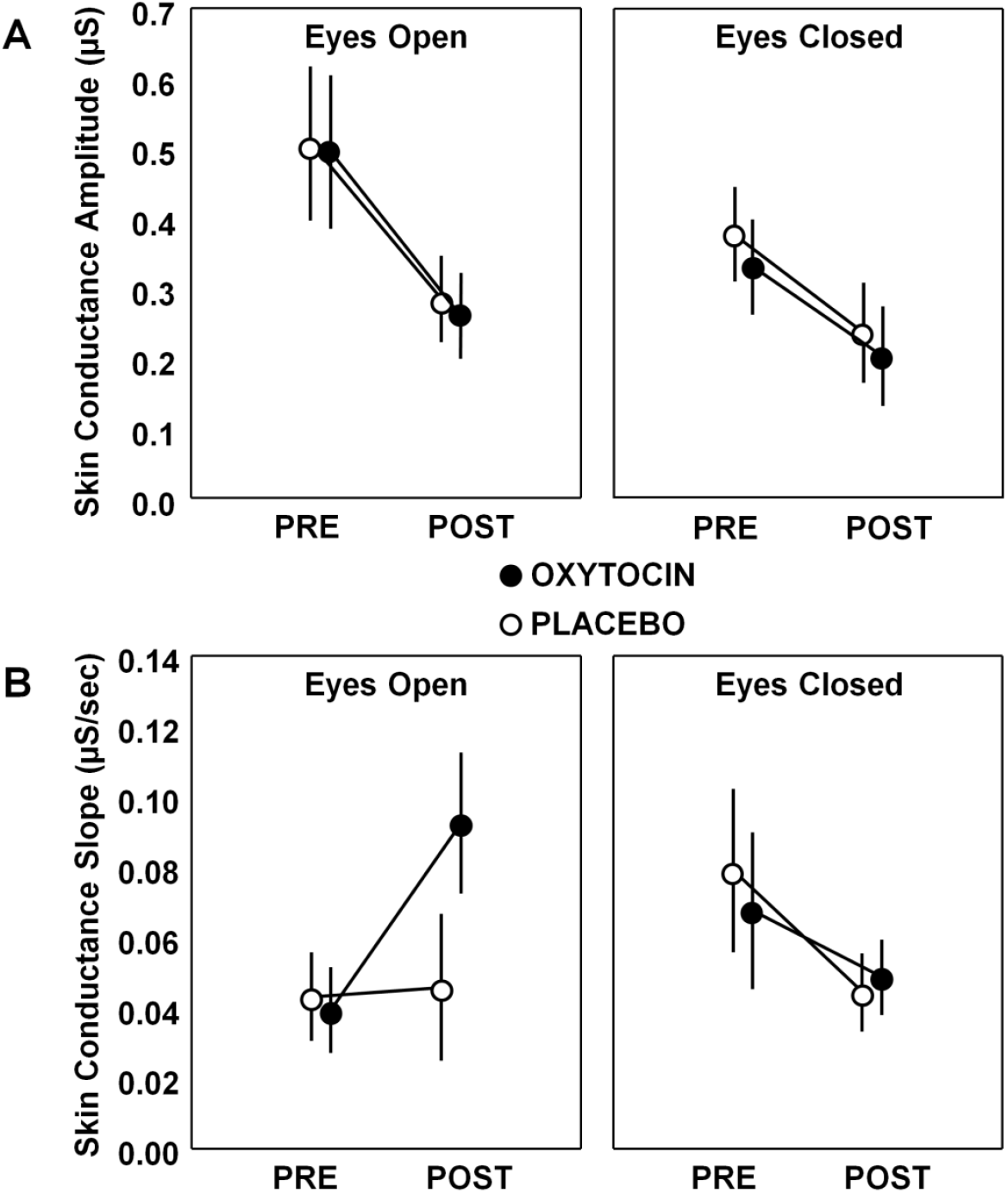
Effect of oxytocin on skin conductance. For each treatment group (oxytocin, placebo), skin conductance amplitudes (**A**) and slopes (**B**) are visualized separately for each condition (eyes open, eyes closed) and assessment session (pre, post). Vertical bars denote +/- standard errors.

**Table 1.** Effects of oxytocin administration. Mean pre-to-post change scores are listed separately for each treatment group (oxytocin, placebo). T and p values correspond to single-sample t tests assessing within-group pre-to-post changes separately within the oxytocin and placebo group. F and p values correspond to between-group differences in pre-to-post change scores (ANOVA ‘time x treatment’ interaction).

| Outcome measure | Oxytocin |  |  |  | Placebo |  |  |  | Between-group difference |  |
| --- | --- | --- | --- | --- | --- | --- | --- | --- | --- | --- |
|  | *N* | *Mean ± SD* | *T value* | *p* | *N* | *Mean ± SD* | *T value* | *p* | *F value* | *p* |
| **Skin conductance response (SCR)** |  |  |  |  |  |  |  |  |  |  |
| Amplitude - eyes open | 25 | -0.23 ± 0.10 | -2.35 | 0.03 | 25 | -0.22 ± 0.11 | -2.02 | 0.05 | 0.01 | 0.93 |
| Amplitude - eyes closed | 25 | -0.13 ± 0.05 | -2.37 | 0.03 | 25 | -0.14 ± 0.10 | -1.46 | 0.16 | 0.01 | 0.91 |
| Slope - eyes open | 25 | 0.05 ± 0.02 | 2.66 | 0.01 | 23 | 0.003 ± 0.01 | 0.25 | 0.80 | **5.15** | **0.028\*** |
| Slope - eyes closed | 25 | -0.02 ± 0.02 | -1.38 | 0.18 | 23 | -0.03 ± 0.02 | -1.53 | 0.14 | 0.34 | 0.56 |
| **Profile of Mood State (POMS)** |  |  |  |  |  |  |  |  |  |  |
| Tension | 28 | -2.43 ± 0.56 | -4.32 | 0.0002 | 25 | -0.76 ± 0.54 | -1.40 | 0.17 | **4.51** | **0.038\*** |
| Anger | 28 | -1.43 ± 0.42 | -3.43 | 0.002 | 25 | -0.88 ± 0.31 | -2.86 | 0.01 | 1.08 | 0.30 |
| Depression | 28 | -1.68 ± 0.44 | -3.84 | 0.0007 | 25 | -1.52 ± 0.61 | -2.48 | 0.02 | 0.05 | 0.83 |
| Fatigue | 28 | 0.54 ± 0.48 | 1.13 | 0.27 | 25 | 0.76 ± 0.61 | 1.25 | 0.22 | 0.09 | 0.77 |
| Vigor | 28 | -1.43 ± 0.63 | -2.28 | 0.03 | 25 | -2.32 ± 0.61 | -3.78 | 0.0009 | 1.02 | 0.32 |
| **Self-Assessment Manikin (SAM)** |  |  |  |  |  |  |  |  |  |  |
| Affective valence | 27 | -0.52 ± 0.25 | -2.05 | 0.05 | 27 | -0.67 ± 0.24 | -2.73 | 0.01 | 0.18 | 0.68 |
| Arousal | 27 | -0.30 ± 0.31 | -0.95 | 0.35 | 27 | -0.85 ± 0.34 | -2.47 | 0.02 | 1.43 | 0.24 |
| **Situational Self-Awareness Scale (SSAS)** | 27 | -0.54 ± 0.17 | -3.23 | 0.003 | 27 | -0.12 ± 0.15 | -0.80 | 0.43 | 3.54 | 0.065 |

For SCR amplitude, negative scores indicate reduced sympathetic arousal. For SCR slope, positive scores indicate faster recovery of SCR to baseline. For POMS tension, anger, depression and fatigue; negative scores indicate pre-to-post improvement. For SAM affective valence and arousal, negative scores indicate pre-to-post change to more pleasant/ less arousing ratings. For SSAS, negative scores indicate pre-to-post reduction in self-awareness.

#### Skin conductance response - slope

ANOVA analysis of the SCR-slope data revealed a significant ‘time x condition’ interaction (*F*(1, 45)= 13.50; *p*= .0006), and at trend-level a ‘treatment x time x condition’ interaction (*F*(1, 45)= 3.55; *p*= .065). Further exploration of the three-way interaction revealed a significant ‘treatment x time’ interaction for the direct eye gaze condition (*F*(1, 46)= 5.15; *p*= .028; *η*^*2*^= .10) (Mann-Whitney U including outliers: *Z*= 2.12; *p*= .034,) (**Figure 1B & Table 1**), but not for the eyes closed condition (*F*(1, 46)= 0.34; *p*= ..56; *η*^*2*^= .007) (Mann-Whitney U test including outliers: *Z*= .12; *p*= .91). As such, compared to PL, OT administration induced an increase in the steepness of slope, indicating a faster recovery (steeper decline) of the eye-contact induced SCR back to baseline (**Figure 1B**).

### Effect of IN-OT on self-rated mood, public self-awareness, arousal and valence

A significant effect of treatment was identified for the mood state ‘tension’ (assessed with the Profile of Mood States: POMS) (*F*(1, 51)= 4.51; *p*= .038; *η*^*2*^= .08), indicating a reduction in reported feelings of tension in the OT group, compared to the PL group (**Figure 2A**). No significant effect of treatment was identified for the other POMS mood states (depression, anger, fatigue, vigor) (all, p > .30) (see **Table 1**).

**Figure 2.**
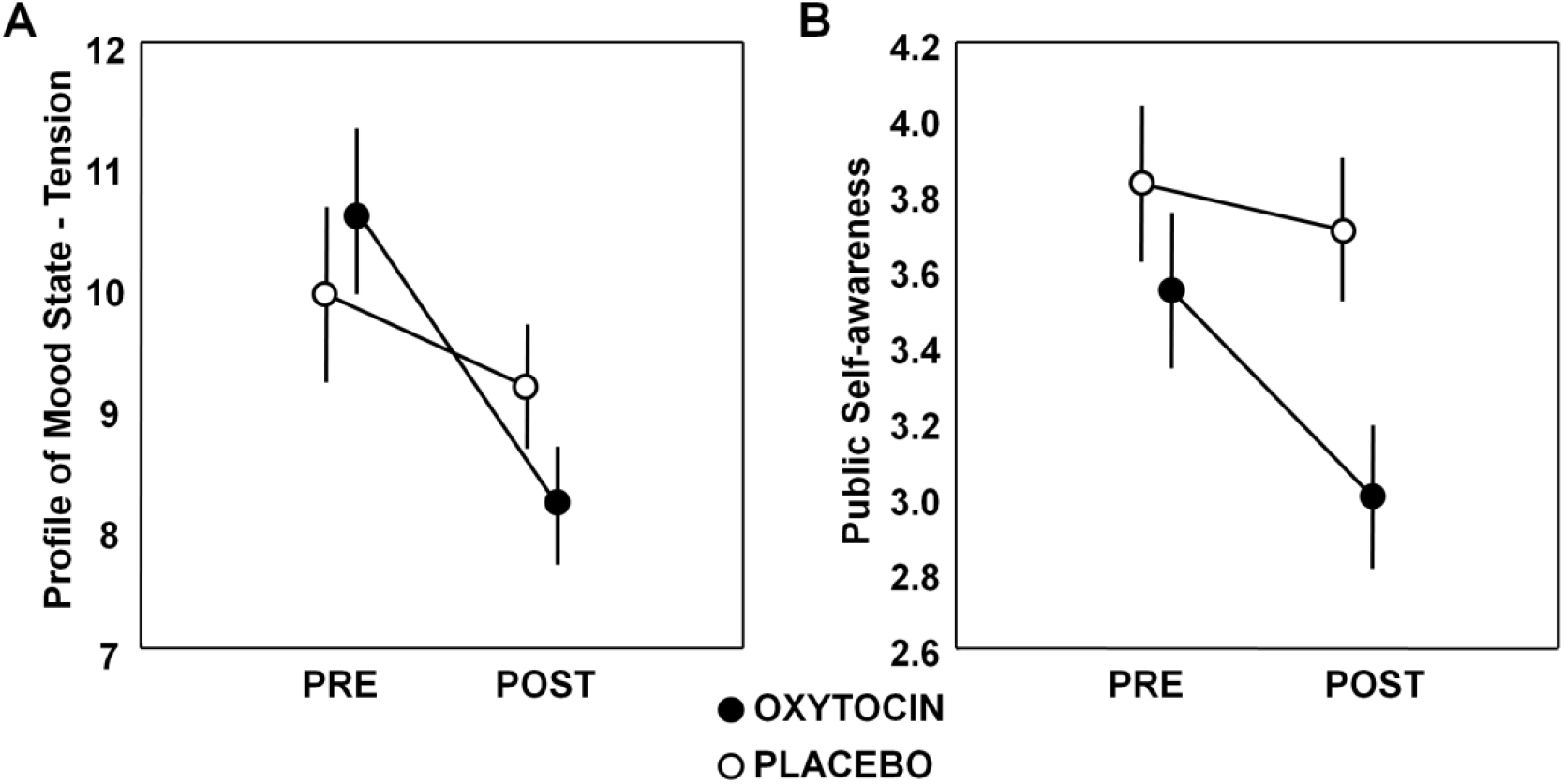
Effect of oxytocin on self-rated mood and public self-awareness. For each treatment group (oxytocin, placebo), self-rated ‘tension’ (assessed with the Profile of Mood States: POMS) (**A**) and public self-awareness (assessed with the Situational Self-Awareness Scale: SSAS) (**B**) are visualized separately for each assessment session (pre, post). Vertical bars denote +/- standard errors.

An effect of treatment was evident at trend-level in terms of public self-awareness (assessed with the Situational Self-Awareness Scale: SSAS) (*F*(1, 52)= 3.54; *p*= .065; *η*^*2*^= .06), indicating a relative pre-to-post reduction in self-awareness (less concerned about themselves) after OT administration, compared to PL administration (**Figure 2B**). No treatment-specific changes were evident in terms of self-rated arousal and ratings of valence (pleasantness) (all, *p* > .24) (see **Table 1**).

### Modulation by variations in person-dependent factors

The effect of IN-OT treatment on SCR-slope was modulated by person-dependent factors, indicating that participants of the IN-OT group with higher self-reported scores on the Social Responsiveness Scale (SRS) (more impairment) showed a stronger treatment-induced increase in SCR-slope (faster recovery) (Spearman *R*_*(25)*_ = .55; *p*= .005). Note that the relationship became more pronounced after removal of two influential data points (Cook’s distance > 3.5) (Spearman *R*_*(23)*_ = .64; *p*= .001) (**Figure 3A**). Participants with high SRS-scores also showed a more pronounced improvement in self-reported public self-awareness after IN-OT (less concerned about themselves) (Spearman *R*_*(27)*_ = -.52; *p*= .006) (**Figure 3B**).

**Figure 3.**
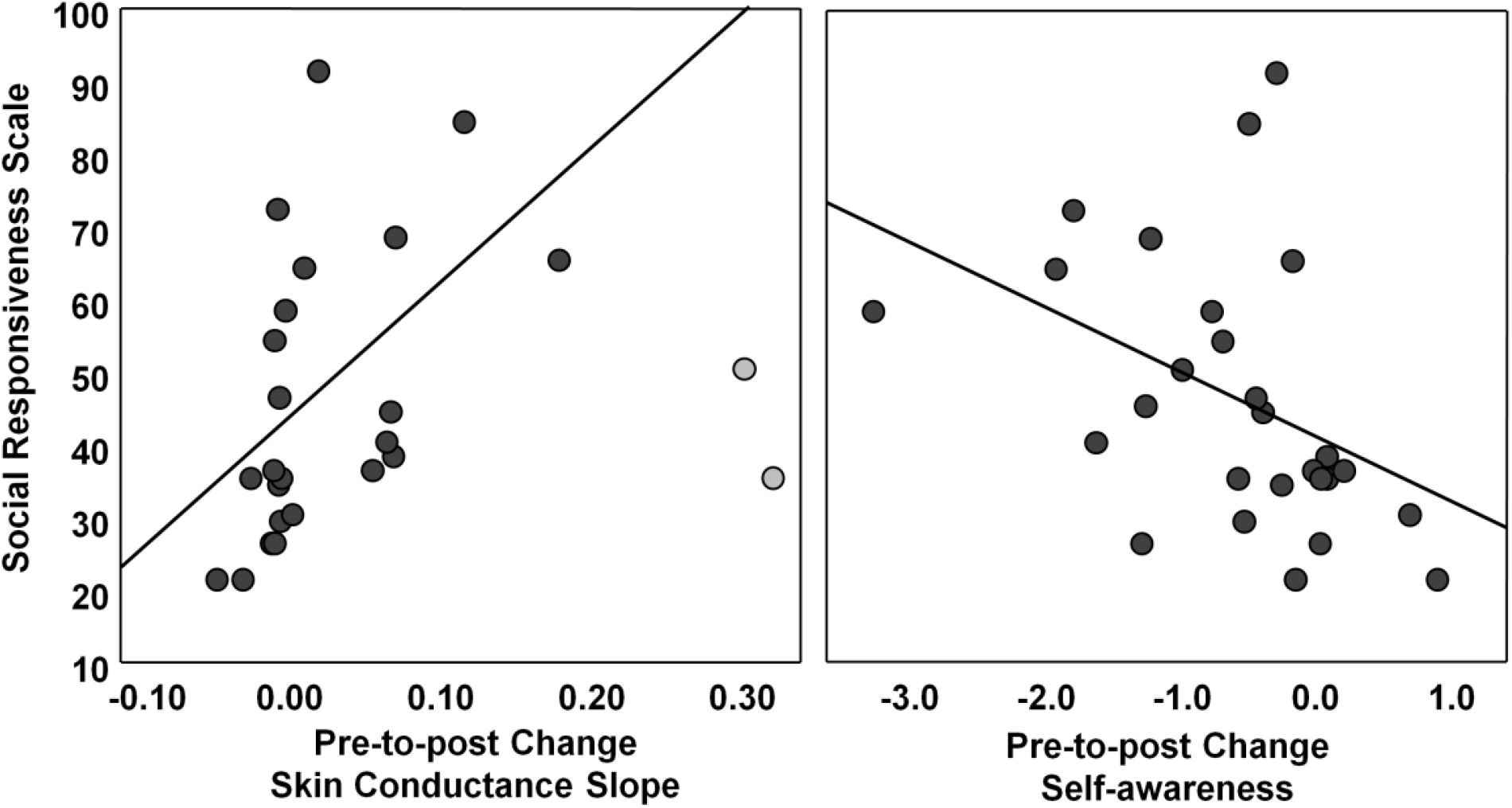
Modulation by person-dependent factors. Visualization of the relationship between scores on the Social Responsiveness Scale (SRS, self-report) and oxytocin-induced pre-to-post changes in skin conductance slopes and public self-awareness (assessed with the Situational Self-Awareness Scale: SSAS). Participants with higher self-reported SRS-scores (more impairment) showed a stronger treatment-induced increase in skin conductance slopes (faster recovery), as well as a stronger reduction in self-awareness (less concerned about themselves). The light-colored data points refer to two influential data points (Cook’s distance > 3.5).

A tentative association was also revealed between attachment avoidance (SAAM) and the treatment-effect on SCR-slope, indicating that participants with higher self-reported attachment avoidance showed a stronger treatment-induced increase in SCR-slope (Spearman *R*_*(25)*_ = .36; *p*= .08). The relationship reached significance (Spearman *R*_*(23)*_ = .52; *p*= .01) after removal of the two influential data points. The treatment-effect on SCR-slope was not significantly modulated by inter-individual variance in SAAM attachment security or anxiety (both, *p* > .20).

## Discussion

Autonomic arousal upon direct eye contact was shown to be modulated by IN-OT, indicating a faster recovery of skin conductance responses (SCR) back to baseline (increased steepness of slope), but no treatment-specific effect of the evoked SCR amplitude per se. The treatment effect on SCR recovery was paralleled by a relative pre-to-post reduction in self-rated feelings of public awareness. Both of these treatment-effects were most prominently observed in participants with low self-reported social responsiveness (higher total scores on the Social Responsiveness Scale; SRS). IN-OT was also shown to induce a general reduction in feelings of ‘tension’ (Profile of Mood States; POMS).

Prior IN-OT studies consistently demonstrated reductions in tonic skin conductance levels during a public speaking test (de Oliveira et al., 2012) and during personal combat imagery (in Vietnam Veterans) (Pitman et al., 1993). In terms of IN-OT effects on phasic skin conductance responses, a more mixed pattern of results was revealed; with one study reporting decreases in SCRs (during late phases of fear extinction (Eckstein et al., 2015)), while other studies reported increases in SCRs (during the acquisition phase of fear conditioning (Eckstein et al., 2016)), or no significant changes in SCRs (during a facial emotion classification task) (Gamer & Buchel, 2012). While direct comparisons between studies are difficult, the present identification of no significant IN-OT effects on SCR-amplitudes upon presentation of direct eye contact stimuli (presented using a live model), seems to be largely in line with the prior study by Gamer et al. (2012), demonstrating no effect of IN-OT on SCR-amplitudes during a facial emotion task (presented with static pictures). The present study included an additional assessment of the effect of IN-OT on the recovery of induced SCRs, and identified that the steepness of negative slope was enhanced after IN-OT, indicating a faster recovery back to initial baseline levels. The obtained pattern of results can be interpreted within the proposed dual action account of OT, implying both a top-down social salience effect (facilitating attention to, and perception of social signals), combined with a bottom-up anxiolytic effect. Firstly, the lack of a treatment-specific effect on SCR-amplitudes may imply that the overall attention allocation to and perception of socially-salient signals such as the eye contact cue was not specifically diminished (or augmented) after IN-OT. However, the identification of a significant IN-OT on SCR recovery provides initial indications that IN-OT may primarily promote the restoration of ‘stressor-induced’ arousal responses, rather than impacting on the overall magnitude of evoked arousal responses per se. Previously, Keri et al. (2011) showed that increases in endogenous OT release during a trust-related social situation (measured in blood plasma) enhanced the habituation of SCRs to a series of auditory tones (Keri & Kiss, 2011). Rodent research showed that both OT and its closely related neuropeptide vasopressin (VP) play an important antagonistic role in alertness and homeostasis, indicating that while VP primarily increases alert and sympathetic drive, OT may primarily decrease alert and enhance parasympathetic outflow (Stoop, 2012). In line with this notion, the present pattern of results provides indications that OT may not directly impact sympathetic arousal responses per se, but instead facilitates recovery to initial baseline arousal states, presumably by enhancing autonomic homeostatic output after (stress-induced) perturbation.

In line with OT’s bottom-up anxiolytic action, the treatment effect on SCR recovery was paralleled by a relative pre-to-post reduction in self-rated feelings of public awareness and feelings of tension. A previous study from our lab (Bernaerts et al., 2017) also adopted the POMS to evaluate the effects of a two-week multiple-dose IN-OT treatment in neurotypical men, and similarly showed OT-specific reductions in feelings of tension (and also anger). The POMS was also adopted in a recent study evaluating the effects of a four-week IN-OT treatment in adult men with autism spectrum disorder (Bernaerts, Boets, Bosmans, Steyaert, & Alaerts, 2020), and while here, no treatment-specific changes in tension were detected (both groups reported improvements), the POMS revealed OT-specific reductions in feelings of vigor (feeling more “active”, “energetic”, “lively”).

Notably, the treatment effect on SCR recovery (and public self-awareness) was most prominently observed in participants with low self-reported social responsiveness (higher total scores on the SRS) and high reports of avoidant attachment (SAAM). These observations are generally in line with the notion that IN-OT treatment responses may be more pronounced for individuals with low baseline levels of social proficiency or approach motivation (e.g. avoidantly attached individuals), whereas for individuals with already high baseline levels of approach motivational tendencies (e.g. securely attached individuals), the additional administration of IN-OT may not stimulate prosocial behavior further (Bartz et al., 2015). In line with the current observation of a modulatory impact of attachment avoidance, two prior studies of our lab also identified SAAM attachment avoidance to be a sensitive modulator of IN-OT treatment responses (Bernaerts et al., 2017; Prinsen et al., 2018). In one study, transcranial magnetic stimulation was adopted to assess automatic motor simulation (mirroring) of others actions and showed that, in avoidantly attached individuals - who otherwise show a low propensity to mirror - a single-dose of IN-OT was able to specifically enhance automatic motor simulation (Prinsen et al., 2018). In the other study, young adult men were administered with a daily dose of OXT for a period of two weeks, and significant improvements in self-reports of attachment avoidance (SAAM) and attachment toward peers were revealed (Bernaerts et al., 2017). Interestingly, and similar to the present study, the treatment-induced changes in the latter study were also found to be most pronounced for participants with less secure attachments. Also, earlier studies by De Dreu et al (2012) showed that IN-OT administration significantly improved cooperation behavior, but particularly in individuals scoring high on attachment avoidance. Furthermore, in terms of social-cognitive competences, Bartz et al. (2010 & 2019) also showed that OT was able to improve empathic accuracy on an emotion recognition task, but only for less-socially proficient individuals (Bartz, Nitschke, Krol, & Tellier, 2019; Bartz et al., 2010). Together, these previous studies and our study add to the growing body of evidence that inter-individual differences in (baseline) person-dependent factors may play a pivotal role in determining IN-OT treatment responses.

While the current study provides important new insights into the effect of IN-OT on eye-contact induced autonomic arousal, several limitations need to be considered. First, since only neurotypical men were included, the current observations of beneficial effects on SCR recovery, self-awareness and feelings of tension cannot be generalized to women. Especially with regard to the adopted live eye gaze stimuli, this is an important issue, considering that prior studies showed that both the participant’s and the model’s gender may influence gaze processing (Ponkanen et al., 2011; Slepian, Weisbuch, Adams, & Ambady, 2011). For example, Pönkänen et al. (2011) showed that for female participants, a significant effect of eye contact on autonomic arousal was only evident for viewing female, but not male faces. Since the investigation of gender differences was beyond the scope of the present study, future research is warranted to systematically assess whether gender (of the participant and model) impacts the effect of IN-OT on eye-contact induced arousal. Future research is also needed to explore whether similar effects are evident in patient populations with particular difficulties in the social domain, such as individuals with autism spectrum disorder or social anxiety disorders.

To conclude, while IN-OT did not specifically enhance or dampen the overall magnitude (amplitude) of sympathetic arousal responses, administration of OT was shown to facilitate the recovery of skin conductance responses back to baseline (increased steepness of recovery slope). Together, these observations provide important evidence that OT plays a pivotal role in modulating autonomic arousal responses, primarily by facilitating restoration of homeostatic balance after (social) stress-induced perturbation.

## Data Availability

Data available upon request.

## Author Disclosures

### Contributions

Author KA designed the study together with author JP. Author KA and ND managed the literature searches and data collection/ processing. Authors KA, ND and JRS undertook the statistical analysis, and author KA wrote the first draft of the manuscript. All authors contributed to and have approved the final manuscript.

## Acknowledgements

The authors would like to thank Elisa Maes for her contribution in conducting the experiment. We would also like to thank all participating subjects and students for their help in data collection.

## Supplementary Information

**Supplementary Table 1.**
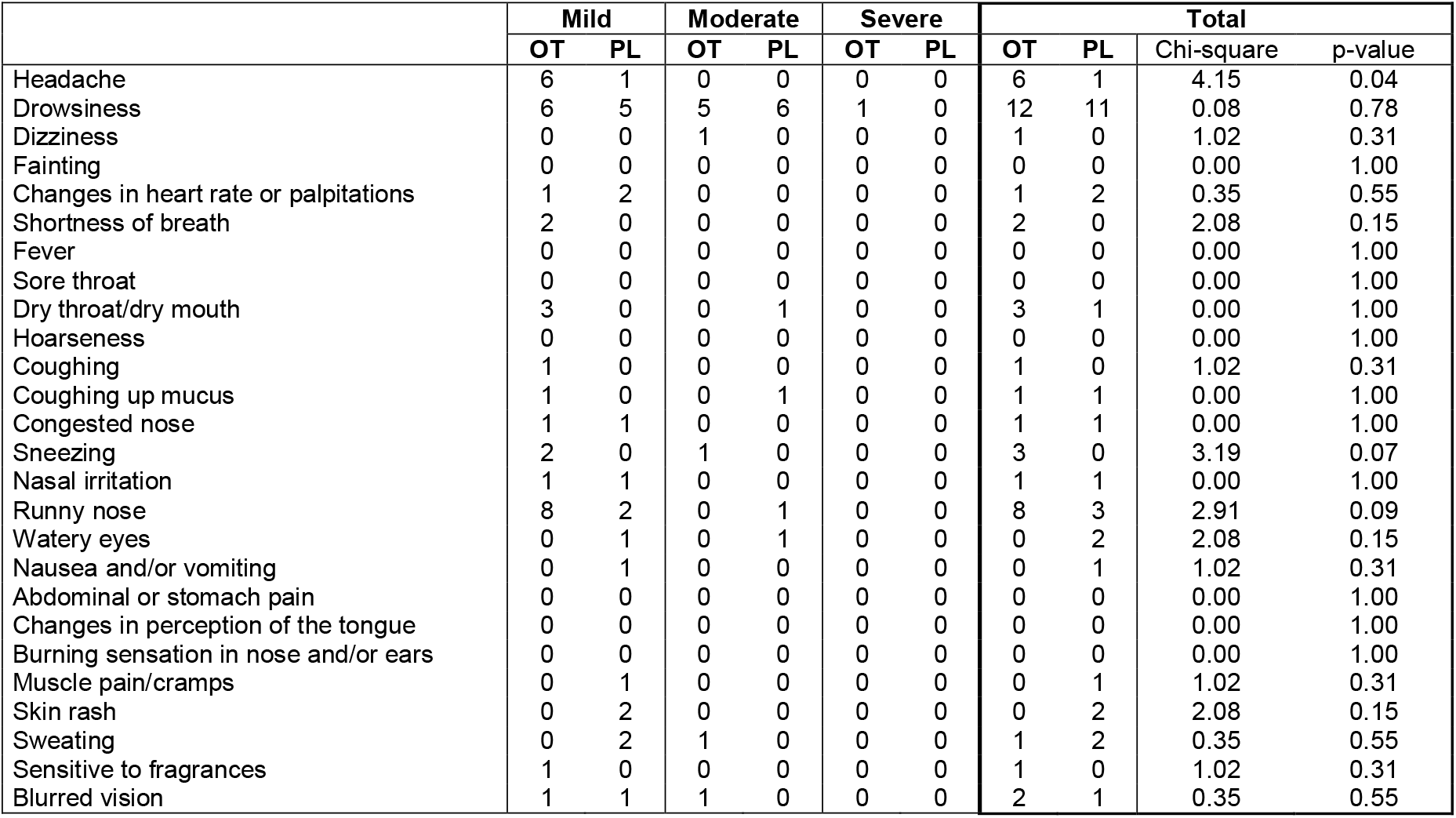
At the end of the experimental session, participants were asked to report whether they presented any of the listed (or other) side effects and to indicate the severity of the side effect (mild, moderate, or severe). The number of OT participants (out of n=25) or PL participants (out of n=25) that reported any mild, moderate or severe side effects are listed separately for each side effect. A significant group difference (Pearson Chi-square test) was noted for the side effect ‘headache’, indicating that a larger number of OT participants reported a (mild) headache.

